# Harnessing recombinase polymerase amplification for rapid detection of SARS-CoV-2 in resource-limited settings

**DOI:** 10.1101/2021.02.17.21251732

**Authors:** Dounia Cherkaoui, Da Huang, Benjamin S. Miller, Rachel A. McKendry

## Abstract

The COVID-19 pandemic has challenged testing capacity worldwide. The mass testing needed to stop the spread of the virus requires new molecular diagnostic tests that are faster and with reduced equipment requirement, but as sensitive as the current gold standard protocols based on polymerase chain reaction.

We developed a fast (25-35 minutes) molecular test using reverse transcription recombinase polymerase amplification for simultaneous detection of two conserved regions of the virus, targeting the E and RdRP genes. The diagnostic platform offers two complementary detection methods: real-time fluorescence or visual dipstick.

The analytical sensitivity of the test by real-time fluorescence was 9.5 (95% CI: 7.0-18) RNA copies per reaction for the E gene and 17 (95% CI: 11-93) RNA copies per reaction for the RdRP gene. The analytical sensitivity for the dipstick readout was 130 (95% CI: 82-500) RNA copies per reaction. The assay showed high specificity with both detection methods when tested against common seasonal coronaviruses, SARS-CoV and MERS-CoV model samples. The dipstick readout demonstrated potential for point-of-care testing, with simple or equipment-free incubation methods and a user-friendly prototype smartphone application was proposed with data capture and connectivity.

This ultrasensitive molecular test offers valuable advantages with a swift time-to-result and it requires minimal laboratory equipment compared to current gold standard assays. These features render this diagnostic platform more suitable for decentralised molecular testing.

## Introduction

At the end of December 2019, a public health alert was released from Wuhan, Hubei province, in China reporting cases of “viral pneumonia of unknown cause” observed in several patients with severe acute respiratory syndrome^1^. Eventually, the newly identified virus was designated as severe acute respiratory syndrome coronavirus 2 (SARS-CoV-2)^2^ and the disease caused by the virus was named COVID-19^3^. As of 8^th^ January 2021, a year after the discovery of the human coronavirus SARS-CoV-2, the World Health Organization reported globally over 86.4 million confirmed cases and 1.8 million deaths from COVID-19^4^.

Rapid development of diagnostic tests for detection of SARS-CoV-2 has been vital to limit the spread of the virus^5^. Molecular diagnosis is necessary to identify patients actively infected when COVID-19 symptoms are not clearly differentiable from other coronaviruses, for instance HCoV-NL63, HCoV-OC43 and HCoV-229E, causing common cold, or the deadly Severe Acute Respiratory Syndrome Coronavirus (SARS-CoV) and Middle East Respiratory Syndrome Coronavirus (MERS-CoV). These tests are also needed to identify asymptomatic cases^6^ (not showing symptoms) or pre-symptomatic cases^7^ (not showing symptoms at the time of test but developing symptoms later) which can be infectious. Since the early stages of the pandemic, the World Health Organization recommended the use of quantitative reverse transcription polymerase chain reaction (qRT-PCR) for nucleic acid amplification as the gold standard diagnostic for SARS-CoV-2^8^. Some in-house qRT-PCR protocols were swiftly developed and recommended for wide use in reference laboratories, such as Hong Kong University (HKU)^9^, Charité Institute of Virology Universitätsmedizin Berlin (Charité-Berlin)^10^ and United States CDC (US CDC)^11^. Although targeting various conserved regions of SARS-CoV-2, these qRT-PCR protocols all function with multiple targets to make the test more sensitive and specific (Supplementary Fig. 1).

Despite the World Health Organisation’s recommendation to use qRT-PCR technology for detection of SARS-CoV-2, the pandemic has highlighted major issues in relying on only one technology: a worldwide shortage of qRT-PCR reagents and instruments considerably slowed down testing^12,13^. The massive number of tests needed to contain the spread of the virus could not be met for many months, even in high-income countries^14^. Having viable alternatives to qRT-PCR for acute COVID-19 that are as sensitive, but faster and simpler to use – particularly in decentralised and resource-limited settings in the low and middle income countries – could increase the testing capacity and reduce community transmission^15^.

Recombinase polymerase amplification (RPA), loop-mediated isothermal amplification (LAMP) and other isothermal amplification methods could meet the needs for mass virological testing as alternatives to PCR^16^. They are portable, faster, giving results in less than 30 minutes compared to several hours with qRT-PCR, enabling testing in settings with scarce resources, without a thermocycler^17,18^. Indeed, the first molecular test to receive FDA authorisation for emergency use, which can be operated by the patient without going to a test centre, harnesses isothermal amplification (RT-LAMP) to deliver an easy-to-use device^19^. Several other RPA-based tests for SARS-CoV-2 reported in the literature present good performances^20,21^.

Reverse transcription RPA (RT-RPA) can rapidly amplify viral RNA and detection of the RPA product has been demonstrated by several methods, the most common are by real-time fluorescence or using dipsticks. An ultrasensitive diagnostic assay using RPA and dipstick was successfully demonstrated by our group for HIV using novel nanoparticles^22^. While LAMP has the advantage of being a patent-free method, it operates at higher temperatures (60-65°C compared to 37-42°C for RPA) and it requires two pairs of primers instead of one, making its design slightly more complex. Choosing an RPA-based amplification enables the reaction to be carried out with basic equipment.

Mobile phone-based diagnostic tests can help reduce the need for specific equipment, whilst providing mobile connectivity, which has emerged as an important criteria in REASSURED diagnostic tests^23^. By taking advantage of their processing speeds, display, storage capacity, their high resolution camera and connectivity, smartphones are useful devices to store information relative to the test and the patient, analyse test results with an enhanced readout compared to the naked eye, to communicate the result to a local hospital and send alerts in case a new outbreak or cluster is detected^24,25^.

Here we report the development of a rapid molecular diagnostic for the detection of SARS-CoV-2 by RT-RPA (Fig. 1a) simultaneously detecting two targets (Fig. 1b). We aimed to design two alternative readouts which are both multiplexed: real-time fluorescence (Fig. 1c) and dipsticks (Fig. 1d). Offering two detection methods makes the assay more accessible to different settings, depending on their resources. The multiplexing of the targets has also been used by gold standard qPCR methods (Supplementary Fig. 1) as it offers a greater confidence on the test result, which can be confirmed if only one gene was detected.

**Fig. 1.**
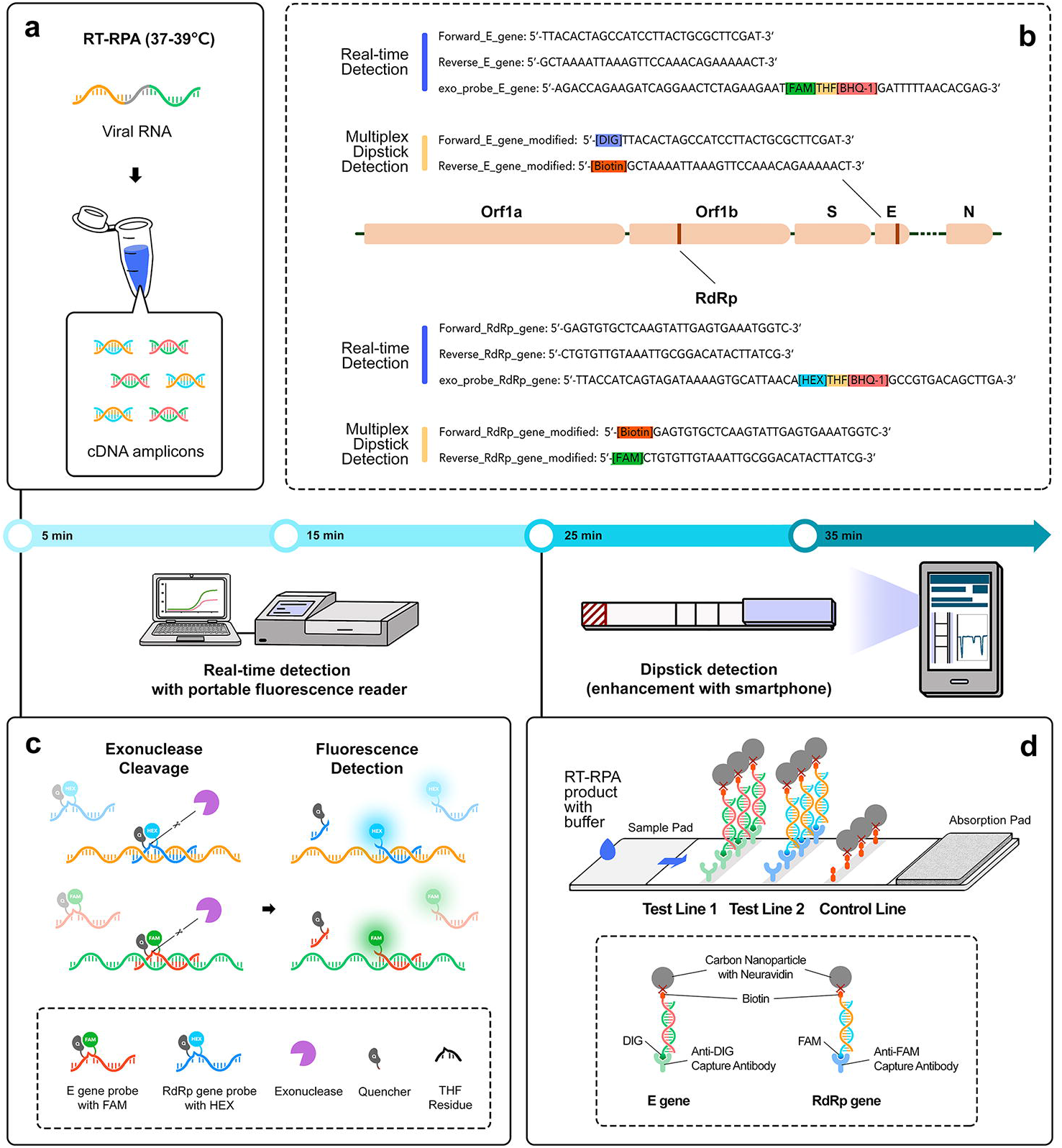
Schematic representation of the rapid and multiplex RT-RPA assay with real-time fluorescence and dipstick detection. a. One-pot RT-RPA assay including reverse transcription of the viral RNA and amplification by RPA at constant temperature (37-39°C). **b** Sequences of the primers/probe sets used for SARS-CoV-2 E gene and RdRP gene in the multiplex RT-RPA assay with real-time detection (blue) and sequences of the modified primers used for the multiplex dipstick detection (orange). **c** Real-time fluorescence detection by exonuclease cleavage of the probes for E gene and RdRP gene at their THF residue. **d** Design of the dipstick for multiplexed detection of the E gene and the RdRP gene.

## Methods

### Reagents and equipment

RPA primers and the cDNA control for the N gene were obtained from Integrated DNA Technologies. The cDNA controls for the E, RdRP and Orf1ab genes were supplied by GenScript Biotech (pUC57-2019-nCoV-PC:E, pUC57-2019-nCoV-PC:RdRP, pUC57-2019-nCoV-PC:ORF1ab). The coronavirus specificity panel (SARS-CoV-2, SARS-CoV, MERS-CoV, HCoV-NL63, HCoV-OC43 and HCoV-229E) was obtained from the European Virus Archive (EVAg). These RNA samples were supplied as full-length virus RNA with reported cycle threshold (Ct) values between 28-30 from qRT-PCR assays. The fluorescent probes were synthesized by Eurogentec. The RPA reactions kits were ordered from TwistDX. The QIAquick Gel Extraction and PCR Purification kits were ordered from Qiagen. The Phusion™ High-Fidelity DNA Polymerase kit and the M-MLV reverse transcriptase were from Thermo Fisher Scientific. The *in vitro* transcription and the RNA purification was performed with the HiScribe™ T7 Quick High Yield RNA Synthesis kit and Monarch RNA CleanUp kit from New England Biolabs Ltd. SUPERase•In™ RNase inhibitor from Invitrogen was added to the RNA standards. Human saliva from healthy and pooled donors (cat. 991-05-P-PreC) was purchased from Lee Biosolutions, Inc. cDNA concentrations were measured on a Nanodrop™ One/One^C^ microvolume UV-Vis spectrophotometer. RNA concentrations were measured on a Qubit 4 fluorometer using the Qubit™ RNA HS Assay Kit (Invitrogen). Fluorescent readings were done on the microplate reader SpectraMax^®^ iD3 from Molecular Devices, for initial screening of SARS-CoV-2 genes, then the Axxin^®^ T16-ISO was used for the duplex diagnostic platform. The dipsticks and running buffer were obtained from Abingdon Health.

### Screening of four different genes (N, E, RdRP and Orf1ab genes) by real-time RPA

RPA primers and probes with a FAM fluorophore were designed for four targets (Supplementary Table 1) and screened using the TwistAmp® exo RPA reactions. The 50 µL reactions contained TwistAmp® exo RPA pellets resuspended in 29.5 µL Rehydration Buffer (TwistDX), 2.1 µL of forward primer (at concentration 10 µM), 2.1 µL of reverse primer (at concentration 10 µM), 0.6 µL of probe (at concentration 10 µM), 1 µL of corresponding cDNA template and 12.2 µL of nuclease-free water. Finally, 2.5 µL of magnesium acetate (at concentration 280 mM) was added to start the reaction. Three cDNA concentrations (50, 500, 5000 copies) were tried along with a non-template control (NTC). The reactions were incubated at 39°C for 30 minutes and real-time fluorescence was recorded using a microplate reader (excitation wavelength 495 nm and emission 520 nm). The screen was done in technical replicates (N=2). Background correction was done on all data by subtracting the measurement value taken at ≈ 60 seconds from all other measurement values, and the values before 60 seconds were set to zero. Then, the average values of duplicates were plotted on GraphPad Prism along with error bars, corresponding to the standard deviation. The fluorescence threshold value for the RPA screen of the four genes with cDNA was set to 25,000. This threshold value was roughly calculated by averaging fluorescence signals from several NTC reactions and adding 3 times the associated standard deviation. The average time to threshold, defined as the time corresponding to the intersection of the amplification curve with the threshold value, was determined for each gene.

### Synthesis of RNA standards for SARS-CoV-2 E and RdRP genes

The plasmid cDNA encoding for the E and RdRP genes were digested using a pair of restriction sites of the plasmid. Double digestion allowed to isolate the sequence of interest and get linear DNA. The product of digestion was run on a 1% agarose gel with a DNA ladder. The band of interest was excised from the gel and the DNA was purified. To generate positive-sense RNA transcripts, a T7 promoter sequence was added via PCR amplification with the promoter sequence on the forward primer (Supplementary Fig. 6a). The PCR products were verified on an agarose gel (Supplementary Fig. 6b). *In vitro* transcription was done with 2.5 hours incubation, with several rounds of DNase I treatment to remove the DNA template, and the RNA was purified. The RNA was tested by PCR using the RPA primers (also suitable for PCR) to check for traces of DNA impurities (Supplementary Fig. 6c). The concentration of the RNA transcripts was measured using the Qubit, then the RNA was diluted in DEPC-treated water and stored at −80°C with RNase inhibitor. A dilution series was used to measure the analytical sensitivity of the molecular test.

### Multiplex RT-RPA with complementary detection platforms

#### Real-time fluorescence readout

Amplification and detection of both genes was done using the multi-channel portable reader (Axxin) using the FAM and HEX channels. The 50 µL reactions contained TwistAmp® exo RPA pellets resuspended in 29.5 µL Rehydration Buffer (TwistDX), 2.1 µL of both forward primers (at concentration 10 µM), 2.1 µL of both reverse primers (at concentration 10 µM), 0.6 µL of both probes, 1 µL of each corresponding RNA samples (E and RdRP genes), 2.5 µL of reverse transcriptase (at 200 U/µL) and 3.9 µL of nuclease-free water. Finally, 2.5 µL of magnesium acetate (at concentration 280 mM) was added to start the reaction. The reactions were incubated at 39°C, with magnetic shaking and the fluorescence was measured in real-time directly from the tubes.

#### Dipstick readout

RPA primers for E and RdRP genes were modified (Supplementary Table 1) for duplex detection on the dipsticks, which incorporate carbon nanoparticles conjugated to neuravidin. The E gene primers were modified with biotin and digoxigenin for detection on test line (1), whereas the RdRP gene primers were modified with FAM and biotin for detection on test line (2). To eliminate non-specific binding due to dimers forming, the assay was tested without any template (negative controls) with modified primers at concentration 10 µM, 2 µM, 1 µM and 0.5 µM (Supplementary Fig. 4). Eventually, the 50 µL reactions contained TwistAmp® basic RPA pellets resuspended in 29.5 µL Rehydration Buffer (TwistDX), 2.1 µL of both forward primer (at concentration 1 µM), 2.1 µL of both reverse primer (at concentration 1 µM), 1 µL of each corresponding RNA sample (E and RdRP genes), 2.5 µL of reverse transcriptase (at 200 U/µL) and 5.1 µL of nuclease-free water. Finally, 2.5 µL of magnesium acetate (at concentration 280 mM) was added to start the reaction. The reactions were incubated at 37°C in an incubator for 20 minutes, with shaking at 250 rpm. Then, 10 µL of reaction was mixed in the well of a microplate with 140 µL of running buffer, and the dipstick was dipped in the well. The test result was read after 10 minutes. A photograph of the strips was taken at this time and image analysis was done on Matlab (R2020b).

Detection of RNA transcripts spiked in human saliva was done following the protocol for RT-RPA with dipstick readout, yet the 5.1 µL of nuclease free-water were replaced by human saliva.

### Calculation of fluorescence thresholds and analytical sensitivity

Two thresholds were calculated for the RT-RPA protocol using the FAM and HEX dyes. The thresholds were computed from eight NTC reactions. The maximum fluorescence values were taken after background subtraction. The average of these values and the standard deviation were calculated. Finally, the thresholds were calculated as followed:

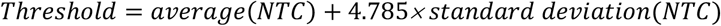

The multiplication factor 4.785 corresponds to the 99.9% confidence interval of the t-distribution with seven degrees of freedom, as per the equation 1 for determining limit of blank^26^. This high confidence interval was chosen to strengthen the specificity of the assay. The analytical sensitivity of the RT-RPA with real-time fluorescence readout was done using these thresholds. Repeats were run five times for a range of RNA inputs: 1, 2.5, 5, 7.5, 10, 10^2^ and 10^3^ (only for the RdRP gene). The fraction of positive reactions (reactions which reached the threshold in less than 20 minutes) was calculated separately for both genes and probit analysis was done on Matlab (R2020b).

The EC_95_ was calculated from the probit analysis with its 95% CI. The EC_95_ was defined as the analytical sensitivity of the test.

### Testing of coronavirus specificity panel

RT-RPA protocol (for both real-time and dipstick readout) was followed to test cross-reactivity of the assay with other coronaviruses, namely SARS-CoV, MERS-CoV, HCoV-NL63, HCoV-OC43 and HCoV-229E. As the RNA concentrations of the stock RNA received from supplier was unknown, the RT-RPA assays were run with 5 µL of RNA directly from stock. The SARS-CoV-2 RNA sample supplied with the specificity panel was also run with 5 µL from stock for comparison with the other coronaviruses.

### Smartphone application

The “CovidApp” smartphone application was developed in Android Studio using java libraries. Screenshots were taken from the emulator using a Galaxy Nexus API 28. The complete code for the Android application is available on request.

The application opens onto a homepage where the users can choose between three activities “Test”, “Alerts” or “Map outbreak”. The “Test” activity includes first recording of patient information (patient ID, date of birth, GPS and symptoms). The GPS coordinates are captured in real-time by clicking the button and time and date are also automatically captured. Then, the user can click on the “Take Test Picture” button to get access to the smartphone camera and take a photograph of the lateral flow test. Manual cropping is required to crop onto the result area of the test (where the lines are). Another activity enables image analysis of the cropped image for enhanced visualisation of the test lines and plotting of the test line intensity. Finally, the user can select between the options “three lines”, “two lines”, “one line”, “no line” which records the test result as “positive”, “presumptive positive”, “negative” or “invalid”. If the user test result is “positive” or “presumptive positive”, the user is taken to the “Contacts” page when clicking on the “Next step” button. This will enable to record the contacts of the positive case so then can be later reached by the local contact tracing system. Finally, the activity “Map outbreak” opens to visualise the location of the tested case on the map. In the “Alerts” activity, the information of the patient, with the test result, can be seen.

### Comparison of four isothermal amplification methods

RT-RPA for visual detection on dipstick was commonly done using a laboratory incubator (New Brunswick(tm) Innova^®^ 42) at 37°C with shaking at 250 rpm. Other incubation methods were tried and compared including incubation in a water bath at 37°C, incubation on a hand warmer bag (HotHands^®^ air activated) releasing heat at ∼36°C after activation and by holding the tube in the hand. The positive and negative reactions using these different incubation methods were all done in parallel with an incubation time of 20 minutes. Then, the reactions were analysed on dipsticks following the dipstick readout protocol.

## Results

### Gene screening for detection of SARS-CoV-2 by real-time fluorescence

A pair of RPA primers and a fluorescent “exo” probe (Supplementary Table 1) were designed to target four conserved regions of the SARS-CoV-2 genome in the nucleocapsid (N) gene, the envelope (E) gene, the RNA-dependent RNA polymerase (RdRP) gene and the open-reading frame 1a/b (Orf1ab). The RPA assay design was optimised for amplicon size of ∼200 bp and long primers of ∼30 bp. Preliminary BLAST analysis indicated that these pairs and probes specifically detect SARS-CoV-2.

A preliminary gene screening aimed to identify the best two primers/probe sets among these four targets, able to achieve rapid and sensitive detection of SARS-CoV-2 in a real-time RPA assay. The gene screening was conducted with cDNA controls rapidly made available by suppliers (Supplementary Fig. 2a). A single fluorescence threshold was used to compare the four targets. All reactions using template, except one (50 copies for the N gene), showed successful amplification of 50, 500 and 5,000 copies with fluorescent signals reaching the threshold in less than 30 minutes (Supplementary Fig. 2b). Then, the average time to threshold was determined for each gene and it was used to compare them (Supplementary Fig. 2c). The two genes with the shortest average time to threshold with 50 copies of cDNA were the E gene, in 14 minutes, and the RdRP gene, in 19 minutes. The Orf1ab gene was slightly slower than RdRP gene, while the N gene showed particularly low sensitivity in the RPA protocol and did not reach the threshold with 50 copies. Eventually, the E and RdRP genes were selected and multiplexed to make an in-house duplex RT-RPA protocol to detect SARS-CoV-2 virus. An analysis of genome variations (determined from 5139 sequenced genomes deposited on https:www.gisaid.org/) for the selected primers and probes confirmed that they target conserved regions of the SARS-CoV-2 genome with low variability comprised between 0.1-0.5% (Supplementary Fig. 3).

### Development of the duplex RT-RPA platforms

The RT-RPA assay was developed with two complementary detection systems (Fig. 1a). First, an optical fluorescent readout similar to qRT-PCR that uses fluorescent probes to monitor real-time amplification of the target was made by multiplexing fluorophores to simultaneously detect the amplicons of the E gene with FAM and RdRP gene with HEX (Fig 1.c). The fluorescent probe was designed as a short ∼45-50 oligonucleotide sequence, complementary to the target sequence. The fluorescent probe included a fluorophore and a proximal quencher, separated by a tetrahydrofuran (THF) residue. When the fluorescent probe recognised the target sequence, it annealed and was cleaved at the THF site by the exonuclease contained in the exo RPA reaction. As the fluorophore was released from its quencher, a fluorescent signal was produced and recorded on a multi-channel and portable fluorescence reader.

A second detection method was developed using dipsticks to detect the amplicons on a nitrocellulose strip using nanoparticle labels. The dipstick-based platform was developed to be as low-cost and minimalist as possible. The primer sequences used were the same as for the real-time fluorescence readout above, but these primers were modified with small molecules to mediate capture of the amplicons on the test lines of the dipstick (Supplementary Table 1). Optimisation of the primer concentration was needed to eliminate non-specific binding on the test lines (Supplementary Fig. 4) attributed to binding of dimerised primers when used in excess (> 1 µM). After the amplification was performed, detection of the two amplicons was possible on two distinct test lines: (1) for the E gene, (2) for the RdRP gene and a control line (C) provided confirmation that the test had worked properly (Fig. 1d).

### Evaluation of the RT-RPA assay with real-time fluorescence detection

The analytical sensitivity was measured for the real-time RT-RPA assay and defined as the concentration of analyte, here synthetic SARS-CoV-2 viral RNA copies per reaction, that can be detected ≥ 95% of the time (< 5% false negative rate).

To determine the analytical sensitivity of the RT-RPA fluorescence readout two thresholds were calculated, to account for the different background fluorescence of the FAM and HEX fluorophores (see Material and methods section). The resulting thresholds were 112 for the E gene (FAM) and 13 for the RdRP gene (HEX). RT-RPA reactions were run for different RNA inputs ranging from 1 copy to 10^5^ copies and real-time fluorescence was recorded. The time to threshold was determined for reactions reaching threshold in 20 minutes of amplification (Fig. 2a). The amplification time was fixed at 20 minutes, as the assay was able to detect as little as 1 RNA. To measure the analytical sensitivity of both genes, we calculated the fraction positive to find and plot the EC_95_ (see “Methods” section). The analytical sensitivity was 9.5 RNA copies per reaction (95% CI: 7.0-18) for the E gene and 17 RNA copies per reaction (95% CI: 11-93) for the RdRP gene (Fig. 2b).

**Fig. 2.**
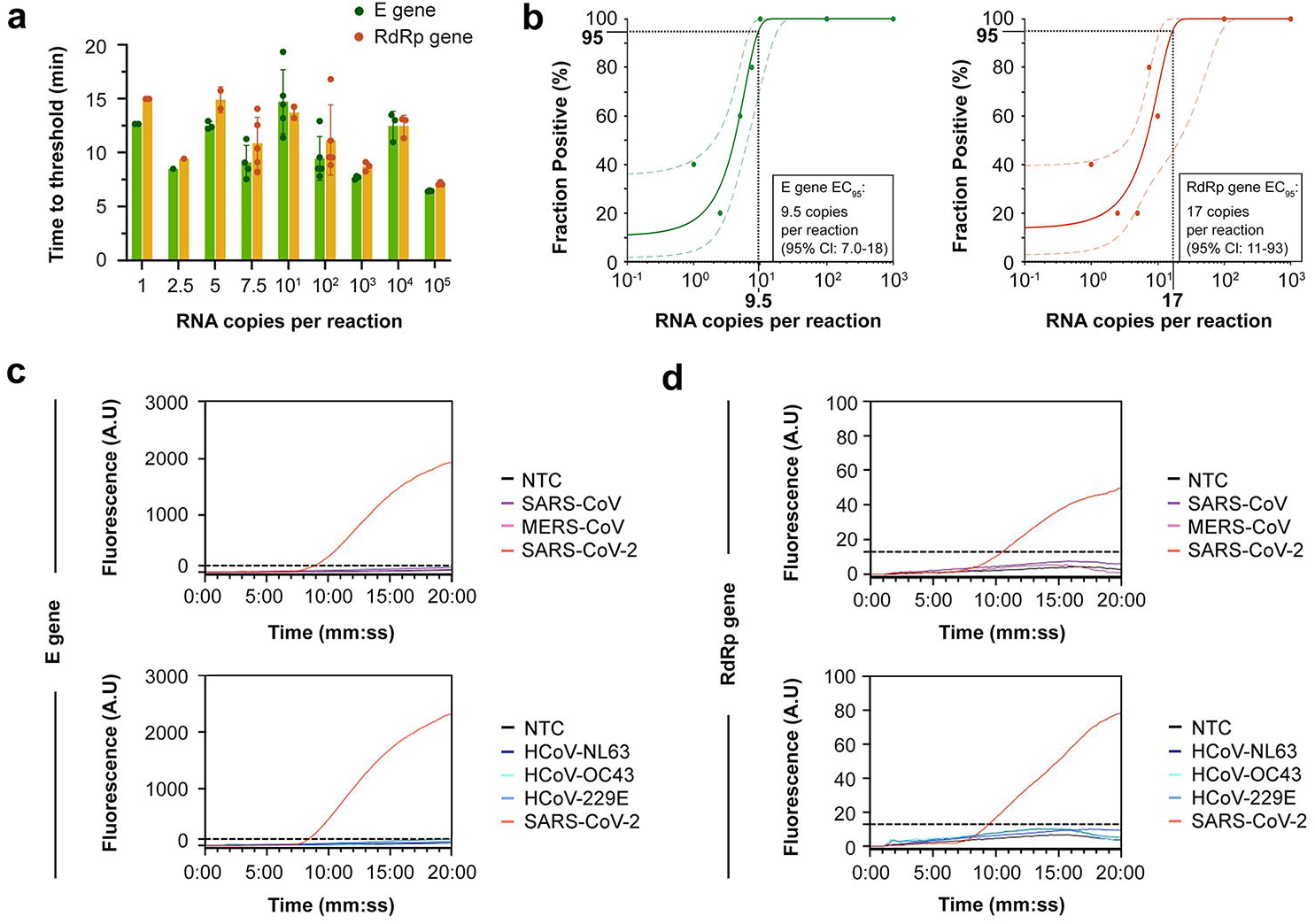
Evaluation of the sensitivity and specificity of the RT-RPA assay with real-time fluorescence detection. **a** Time to threshold for positive RT-RPA reactions with real-time fluorescence detection. The dots represent individual values for the positive reactions reaching threshold for the E gene (in green) and for the RdRP gene (in orange). The bars represent the average time to threshold for the positive reactions and the error bars represent the standard deviation. Each RNA concentration was run in five replicates (N=5), only the positive reactions are represented. **b** Probit analysis for the E gene (left, green) and RdRP gene (right, orange) with their 95% confidence interval (CI). The fraction positive was determined from the RT-RPA reactions in **a** and the probit analysis was done to find the effective concentration at 95% (EC_95_) for both genes. **c** Validation of the specificity of the E gene primers/probe set against SARS-CoV, MERS-CoV (top) and the seasonal coronaviruses (bottom). **d** Validation of the specificity of the RdRP gene primers/probe set against SARS-CoV, MERS-CoV (top) and the seasonal coronaviruses (bottom). NTC: non-template control.

The specificity of the RT-RPA assay was tested with model samples against common seasonal coronaviruses, namely HCoV-NL63, HCoV-OC63 and HCoV-229E, as their symptoms could be easily confused with COVID-19, and we also tested cross-reactivity with SARS-CoV and MERS-CoV, as they are closely related viruses.

No cross-reactivity was observed with the primers/probe set targeting the E gene and the RdRP gene when tested with SARS-CoV- and MERS-CoV and the common colds (Fig. 2c and Fig. 2d). A slight increase in background signal could be observed, although remaining comparable to the non-template control (NTC) reaction and the signal remained below the thresholds.

### Evaluation of the RT-RPA assay with visual dipstick detection

The analytical sensitivity of the dipstick detection method was approximated by running a range of RNA inputs, from 1 to 10^5^ copies. Six replicates were performed (Supplementary Fig. 5) of which one representative dipstick per RNA concentration is shown in Fig. 3a. The test line intensity analysis was used to quantify test line intensity. Single-copy detection was possible for 2/6 repeats (33%), giving in a positive result, defined as both test lines visible by eye or with image analysis. The probit analysis was performed to determine the analytical sensitivity of the assay using the fraction positive (Fig. 3b). The analytical sensitivity of the dipstick method was 130 (95% CI: 82-500) RNA copies per reaction.

**Fig. 3.**
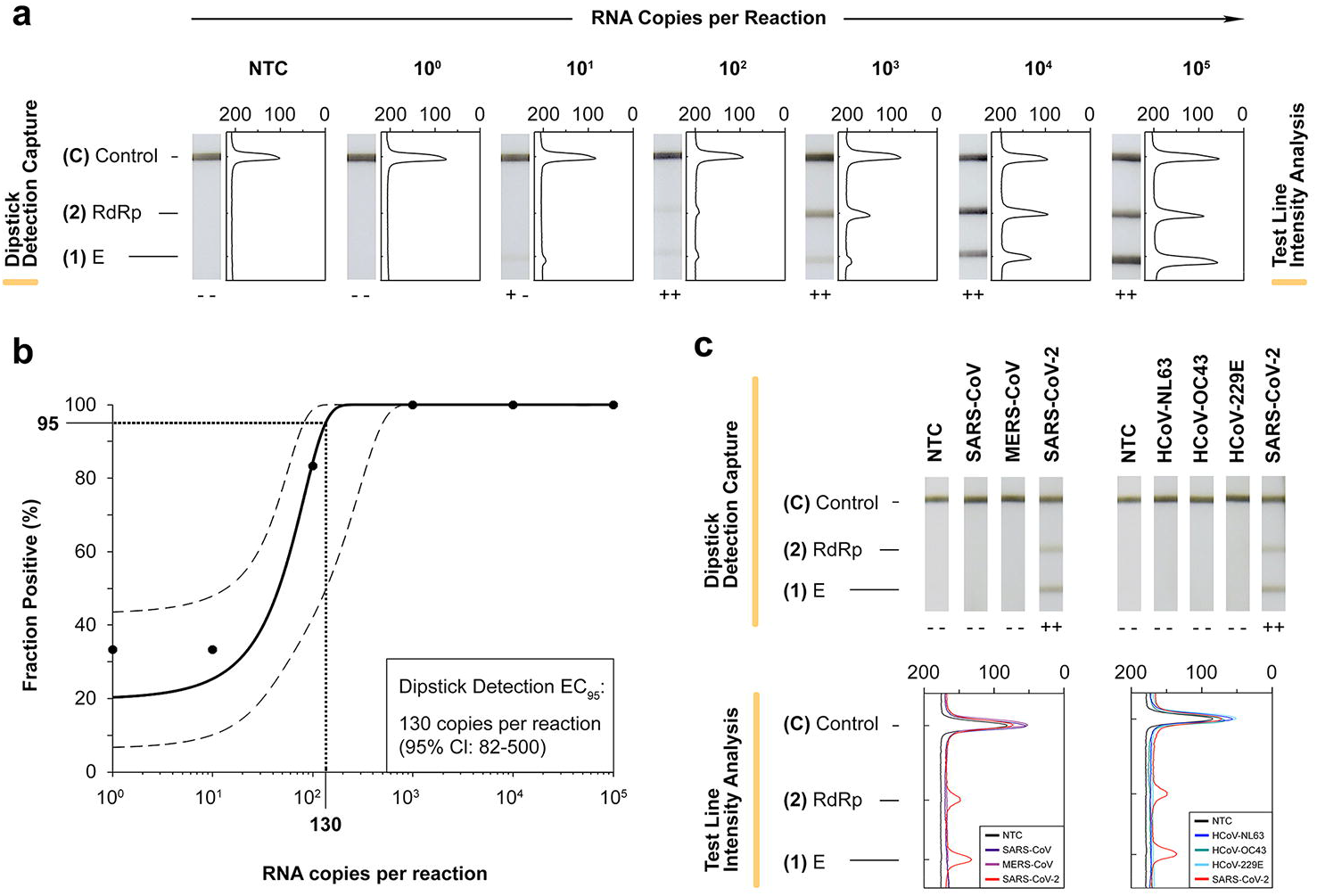
Evaluation of the sensitivity and specificity of the RT-RPA assay with dipstick detection. **a** Evaluation of the sensitivity of the RT-RPA with multiplex dipstick detection. Captures of the dipsticks run with a range of RNA inputs are shown with the associated test line intensity analysis. Dipsticks were annotated (**--**) if no test line was visible (“Negative”), (**+ -**) or (**-+**) if only one test line was visible (“Presumptive positive”) and (**+ +**) if both test lines (1) and (2) were visible (“Positive”). One representative dipstick capture is shown here. **b** Probit analysis and determination of the EC_95_ for the dipstick detection method (taking both genes into account) with the 95% confidence interval (CI). The fraction positive was determined from six replicates (N=6) RT-RPA reactions. **c** Specificity of the dipstick detection method against SARS-CoV, MERS-CoV (left) and the seasonal coronaviruses (right). Photographs of the dipsticks are shown (top) with the associated test line intensity analysis (bottom). NTC: non-template control.

The specificity of the dipstick detection method was assessed against the common seasonal coronaviruses, SARS-CoV and MERS-CoV (Fig. 3c). The dipstick showed high specificity for only SARS-CoV-2 viral RNA and no cross-reactivity was seen with the other coronaviruses.

### Exploration of point-of-care testing with the RT-RPA dipstick method

We investigated the potential of the RT-RPA assay for detection of SARS-CoV-2 at the point-of-care with the dipstick readout, a format that could dramatically widen access to testing in decentralised settings.

The tests could be read visually by eye. In addition, we developed a smartphone application as a prototype towards a connected-diagnostic dipstick test. The architecture and screenshots of the prototype application are presented in Fig. 4a. The smartphone application proposed allowed input and storage of patient’s information, symptoms, capture of geo-location, test lines intensity analysis of the dipstick and a record of the test results. If the test is “Positive” or “Presumptive Positive” the user could insert the names of close contacts for contact tracing purposes. The application also included geographic visualisation of the tested patients to map ‘hotspots’.

**Fig. 4.**
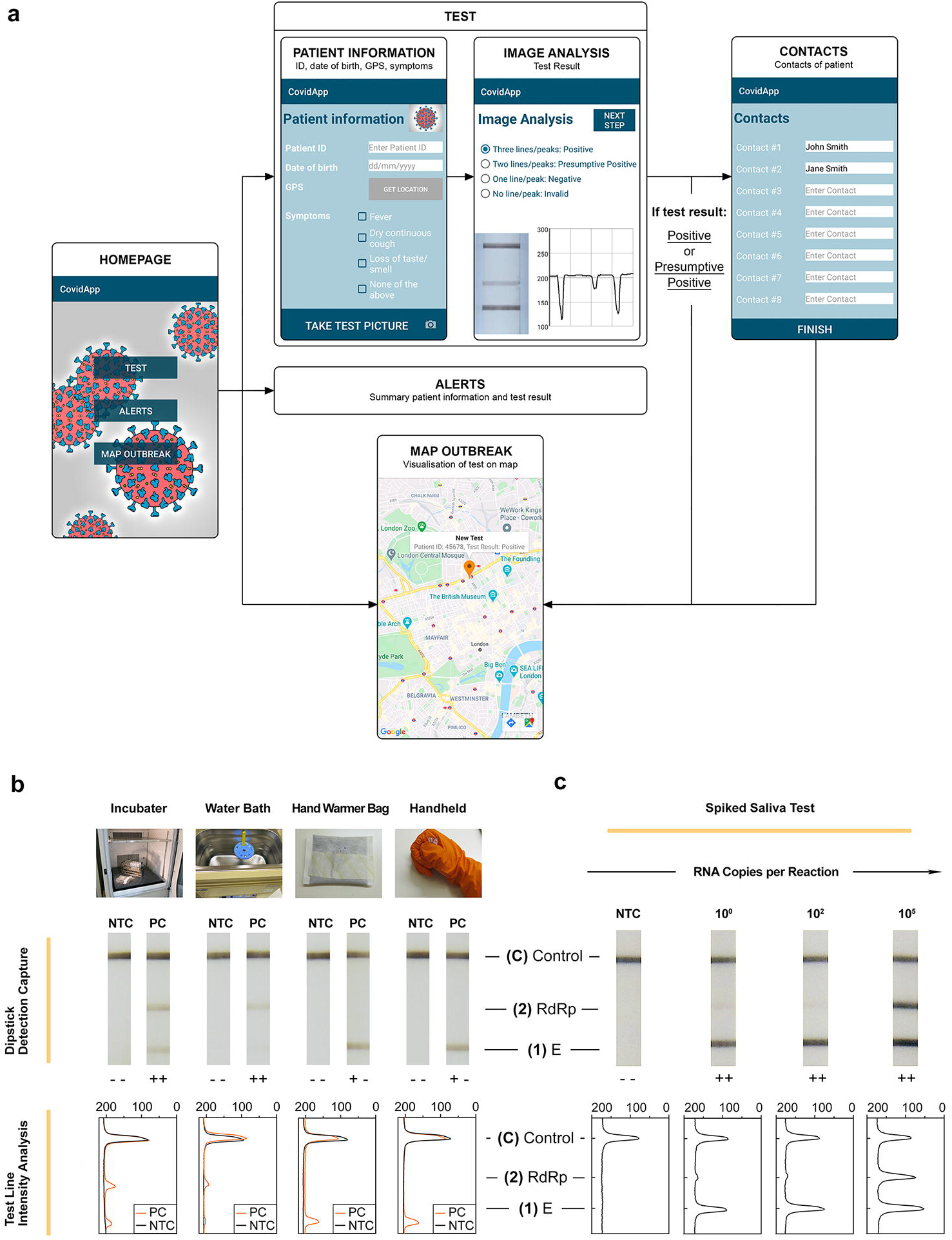
Exploration of point-of-care testing with the RT-RPA dipstick method. a Architecture of the prototype smartphone application “CovidApp”. The design of the smartphone application is represented along with screenshots of the different activities of the application. The main activities, including “Homepage”, “Test”, “Contact”, “Alerts” and “Map Outbreak” are described. b Comparison of incubation of four methods for RT-RPA at ∼37°C with dipstick readout, including incubation using a traditional laboratory incubator with shaking, water bath, hand warmer bag and handheld (using body temperature) (photographs on the top). The lateral flow test captures are shown (middle) with the associated test line intensity analysis (bottom). c Detection of RT-RPA reactions with mock clinical samples (saliva spiked with RNA). Photographs of the lateral flow test captures are shown (top) with the associated test line intensity analysis (bottom). b and c dipsticks were annotated (--) if no test line was visible (“Negative”), (+ -) or (-+) if only one test line was visible (“Presumptive positive”) and (+ +) if both test lines (1) and (2) were visible (“Positive”). NTC: non-template control; PC: positive control (100 copies RNA/reaction).

The major advantage of RPA, compared to other approaches such as PCR and LAMP, is its isothermal amplification at ∼37°C. We investigated the potential of different incubation methods which could be more suitable for point-of-care settings. RT-RPA was performed to detect 100 copies of RNA using four incubation approaches: an incubator, a water bath, a disposable hand warmer bag and simply holding the tube in our hands. Incubators and water baths are often found in well-equipped laboratories, but we also tried using a low-cost hand warmer bag (based on an exothermic reaction shown to deliver a constant temperature of ∼36-37°C for several hours^27^) and holding the tube in one hand (using body temperature ∼37°C) to show inexpensive and equipment-free alternatives. The results are shown in Fig. 4b. While amplification in the incubator seemed to show the best results with two test lines visible on the dipstick, two test lines were also visible for the reaction incubated in the water bath, although slightly fainter. The reactions incubated on a hand warmer bag and handheld appeared less sensitive, showing only a signal on test line (1). However, we proved that very simple methods could be successfully used to amplify SARS-CoV-2 RNA via RT-RPA and visual dipstick detection.

Finally, preliminary analysis was performed to assess the potential of the dipstick test to be compatible with mock clinical samples, using human saliva with spiked RNA transcripts to mimic mouth swabs (Fig. 4c). Saliva is an easy specimen for self-collection that has been FDA-approved for molecular testing of COVID-19^28^. The E gene was clearly detectable on test line (1) with ≥ 1 RNA copy per reaction, and a faint signal was seen on test line (2) for the RdRP gene with 1 and 100 RNA copies per reaction. Two strong test lines were visible for 10^5^ copies per reaction. Therefore, the findings of this small study suggest that conducting the assay in saliva compared to buffer did not have a substantial impact on the assay sensitivity.

## Discussion

Herein we report the development and evaluation of a rapid (25-35 minutes), multiplexed molecular diagnostic for SARS-CoV-2 by RT-RPA. The test was presented with two complementary detection methods: real-time fluorescence using a portable reader and visual dipstick readout on a low-cost nitrocellulose strip. The test showed high sensitivity and high specificity for both readouts. The detection method by dipstick was further investigated for point-of-care and decentralised testing using different incubation methods and a smartphone application to capture, analyse and connect test results.

The development of the multiplex isothermal RT-RPA assay started by selecting two optimal targets, in the E gene and RdRP gene, for rapid and ultrasensitive detection of SARS-CoV-2. Detecting several targets in a multiplex test was done to increase the robustness of the assay, as a common strategy also seen with PCR protocols (Supplementary Fig. 1).

Isothermal fluorescence readers are usually available in centralised laboratories; however, they are not necessarily found in decentralised laboratories and low-resource settings. For this reason, we developed a second readout format, using a dipstick. Dipsticks are portable, cost-effective and user-friendly tools that can detect RPA amplicons with minimal equipment and the test result can be seen with the naked eye. Here only a tube or microplate to mix the RT-RPA reaction with the buffer and a pipette to apply the mix on the dipstick was needed. Giving the option of two alternative readouts with their own advantages aimed to make molecular testing more widely accessible and suitable for decentralised testing.

The amplification time for the RT-RPA assay was set to run for 20 minutes as it was enough to achieve single-copy detection of the E gene with real-time fluorescence and visual dipstick readouts, showing the ultrasensitive potential of the test. The analytical sensitivities for the E and RdRP genes comparable with those reported by Charité for its qRT-PCR assay which were 5.2 copy per reaction (95% CI: 3.7-9.6) for the E gene and 3.8 copy per reaction (95% CI: 2.7-7.6) for the RdRP gene, in comparison to 9.5 RNA copies per reaction (95% CI: 7.0-18) for the E gene and 17 RNA copies per reaction (95% CI: 11-93) for the RdRP gene reported herein. It was necessary to achieve high sensitivity for the molecular test to detect viral loads that are clinically relevant for the disease, here COVID-19. The RT-RPA assay was shown to be highly specific to SARS-CoV-2, with no observed cross-reactivity with the closely related coronaviruses tested, such as SARS-CoV, MERS-CoV, HCoV-NL63, HCoV-OC43 and HCoV-229E. This high specificity was demonstrated for both detection methods and reduces the risk of false positives with closely related viruses.

The prototype smartphone application was proposed as a powerful tool for data capture, analysis and visualisation when testing in decentralised settings. Smartphones are widely accessible, easy-to-use and can act as a substitute to sophisticated laboratory equipment as they integrate a high-resolution camera, large data storage space, real-time location and connectivity. Altogether, these features make health-related smartphone applications attractive accessories to elevate point-of-care diagnostic tests. Moreover, the use of inexpensive methods for incubation at 37°C of the RT-RPA reaction for detection on dipsticks, especially with a hand warmer bag (∼ $0.5) and using body temperature, emphasised the simplicity of the assay.

## Conclusion

To close, we have developed an ultrasensitive and specific diagnostic for SARS-CoV-2 viral RNA using isothermal RPA technology, and proposed two different detection methods, both showing high accuracy. While real-time fluorescence detection developed here offers more sensitivity and faster results (10 minutes faster than dipstick method), the proposed detection on dipsticks appeared as the best method for decentralised testing. Having an alternative to qRT-PCR that is comparable in performance, but with a shorter time-to-result, using different supply chains, requiring less equipment and non-extensive laboratory experience could help to alleviate the pressure on healthcare systems and curb the COVID-19 pandemic worldwide.

Further test development will include clinical validation of the RT-RPA assay with clinical samples with cross-validation of the developed assay with qRT-PCR results to determine the clinical sensitivity and specificity of the test. Preliminary analysis of the dipstick readout with human saliva showed no cross-reaction between mock clinical samples and the RT-RPA assay, hence saliva appeared as a good specimen candidate for a non-invasive test using mouth swab. In future, the adaption of multiplexed gene analysis to detect the S gene could help to track the proportion of new variants of concern and the impact of COVID-19 vaccinations.

## Data Availability

The datasets generated during and/or analysed during the current study, and the computer code used are available from the corresponding author on reasonable request, in line with the requirements of UCL and the funder (EPSRC policy framework on research data).

## Acknowledgments

The authors would like to thank the European Virus Archive (EVAg) for providing the coronavirus specificity panel free of charge. The authors thank Eleanor Gray for her help with synthesis of the RNA standards. The authors also thank Valerian Turbe for his help with the development of the smartphone application. The authors would like to thank Diluka Peiris for support in reagents and equipment acquisition. The authors thank Erin Manning and Jo McHugh for support in communication and project management.

## Ethics statement

Human saliva used as sample was purchased from Lee Biosolutions. The details of the sample, the company and the catalog number can be found in the Material and Methods section of the manuscript. Lee Biosolutions provided an ethics statement confirming that the saliva samples are obtained from anonymised donors in an ethically-sound method and on a voluntary basis.

## Author contributions

Dounia Cherkaoui: Conceptualisation, investigation, methodology, formal analysis, software, project administration, writing – original draft, writing – review & editing.

Da Huang: Conceptualisation, methodology, formal analysis, visualisation, project administration, writing – review & editing

Benjamin S. Miller: Conceptualisation, software, methodology, writing – review & editing Rachel A. McKendry: Conceptualisation, funding acquisition, supervision, writing – review & editing

## Supplementary information

File: Supplementary Table and Figures.docx

