## Supplementary Table and Figures for "Harnessing recombinase polymerase amplification for rapid detection of SARS-CoV-2 in resource-limited settings"

Name: Professor Rachel McKendry

Address: London Centre for Nanotechnology, University College London, 19 Gordon Street, London, United Kingdom

Telephone number: +44 (0)20 7679 9995

**Supplementary Table 1.** RPA primers and exo probes for multi-gene screening of SARS-CoV-2 targets.

| Target | Name | Sequence (5'-3') |
|---|---|---|
| N gene | forward_N_gene | CGCATTACGTTTGGTGGACCCTCAGATTCAACTGG |
| | reverse_N_gene | TTATTGGGTAAACCTTGGGGCCGACGTTGTTTTGA |
| | exo_probe_N_gene (FAM) | CCGACGTTGTTTTGATCGCGCCCCACTGCGT[FAM][THF][BHQ1]CCATTCTGGTTACTGC[Spacer C3] |
| E gene | forward_E_gene | TTACACTAGCCATCCTTACTGCGCTTCGAT |
| | reverse_E_gene | GCTAAAATTAAAGTTCCAAACAGAAAAACT |
| | forward_E_gene_modified | [DIG]TTACACTAGCCATCCTTACTGCGCTTCGAT |
| | reverse_E_gene_modified | [Biotin]GCTAAAATTAAAGTTCCAAACAGAAAAACT |
| | exo_probe_E_gene (FAM) | AGACCAGAAGATCAGGAACTCTAGAAGAAT[FAM][THF][BHQ1]GATTTTTAACACGAG [Spacer C3] |
| RdRP gene | forward_RdRP_gene | GAGTGTGCTCAAGTATTGAGTGAAATGGTC |
| | reverse_RdRP_gene | CTGTGTTGTAAATTGCGGACATACTTATCG |
| | forward_RdRP_gene_modified | [FAM]GAGTGTGCTCAAGTATTGAGTGAAATGGTC |
| | reverse_RdRP_gene_modified | [Biotin]CTGTGTTGTAAATTGCGGACATACTTATCG |
| | exo_probe_RdRP_gene (FAM) | TTACCATCAGTAGATAAAAGTGCATTAACA[FAM][THF][BHQ1]GCCGTGACAGCTTGA[Spacer C3] |
| | exo_probe_RdRP_gene (HEX) | TTACCATCAGTAGATAAAAGTGCATTAACA[HEX][THF][BHQ1]GCCGTGACAGCTTGA[Spacer C3] |
| ORF1ab gene | forward_Orf1ab_gene | AAGGATTTTGTGACTTAAAAGGTAAGTATG |
| | reverse_Orf1ab_gene | GACGGGCTGCACTTACACCGCAAACCCGTT |
| | exo_probe_Orf1ab_gene (FAM) | TGGGTTCGCGGAGTTGATCACAACTACAGC[FAM][THF][BHQ1]AACCTTTCCACATAC [Spacer C3] |

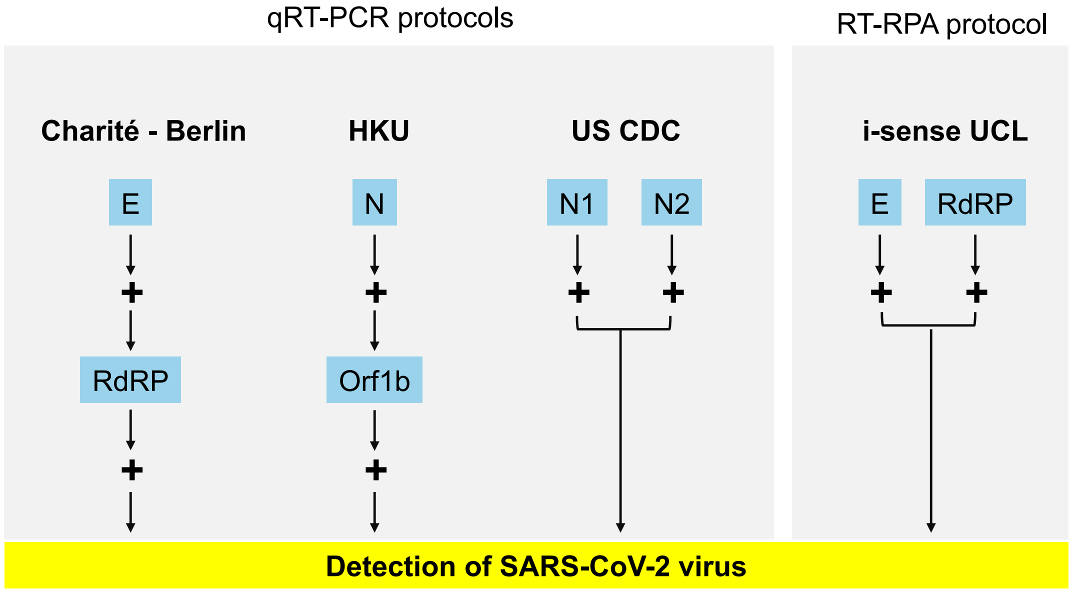


**Supplementary Figure 1.** Comparison of in-house molecular diagnostic qRT-PCR protocols with the developed RT-RPA protocol. Three qRT-PCR protocols developed at Charité Institute of Virology Universitätsmedizin Berlin (Charité-Berlin), Hong Kong University (HKU) and United States CDC (US CDC) and commonly used as gold standard protocols for molecular testing of SARS-CoV-2 were presented with their target genes and workflow.

a

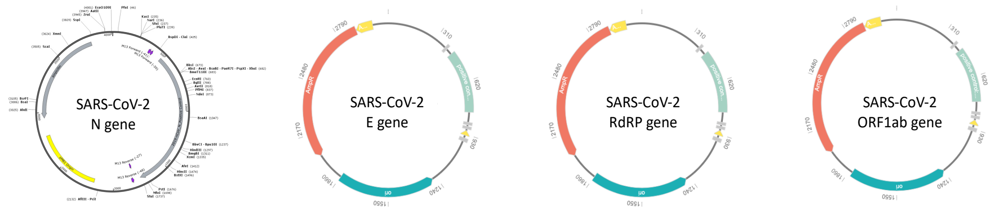


b

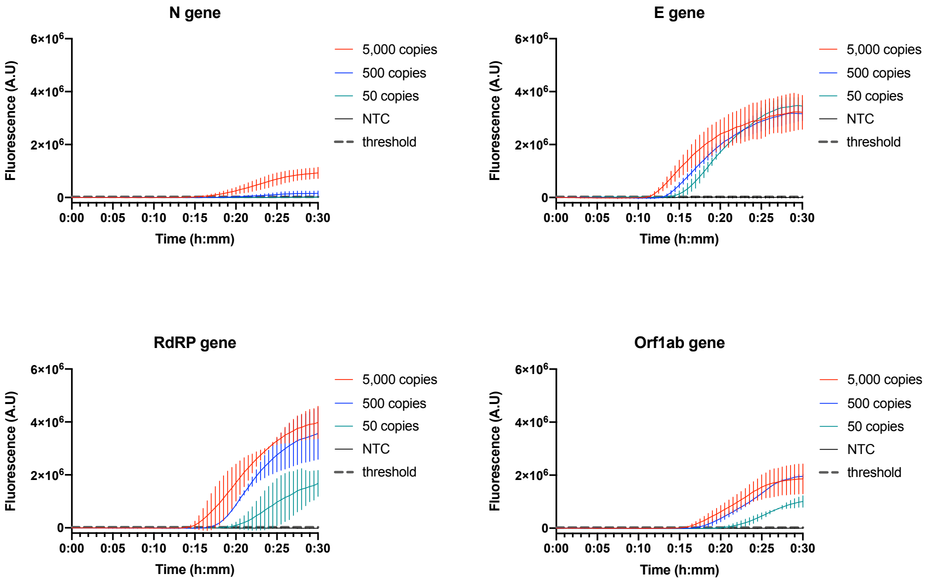


c

| Average time to threshold | | | |
|---|---|---|---|
| **cDNA copies per reaction** | **50** | **500** | **5000** |
| **N gene** | **N\A** | **20:30** | **15:30** |
| **E gene** | **14:00** | **13:30** | **11:30** |
| **RdRP gene** | **19:00** | **17:00** | **14:30** |
| **Orf1ab gene** | **20:30** | **17:00** | **16:00** |

**Supplementary Figure 2.** Gene screening by real-time RPA with control cDNA. (a) Maps of the cDNA control plasmids used for the N gene, E gene, RdRP gene and Orf1ab gene for the screening. Images were provided by the suppliers (N gene: Integrated DNA Technologies; E, RdRP and Orf1ab genes: Eurogentec). (b) Four genes (N, E, RdRP and Orf1ab) were screened by RPA using control cDNA to select the two best targets to be multiplexed in the final RT-RPA assay. Technical replicates (N=2) were averaged and plotted and the error bars represent the standard deviation between replicates. (c) The average "time-to-threshold" was measured and compared for all four genes and the different cDNA inputs (50, 500 or 5,000 copies per reaction).

**a**

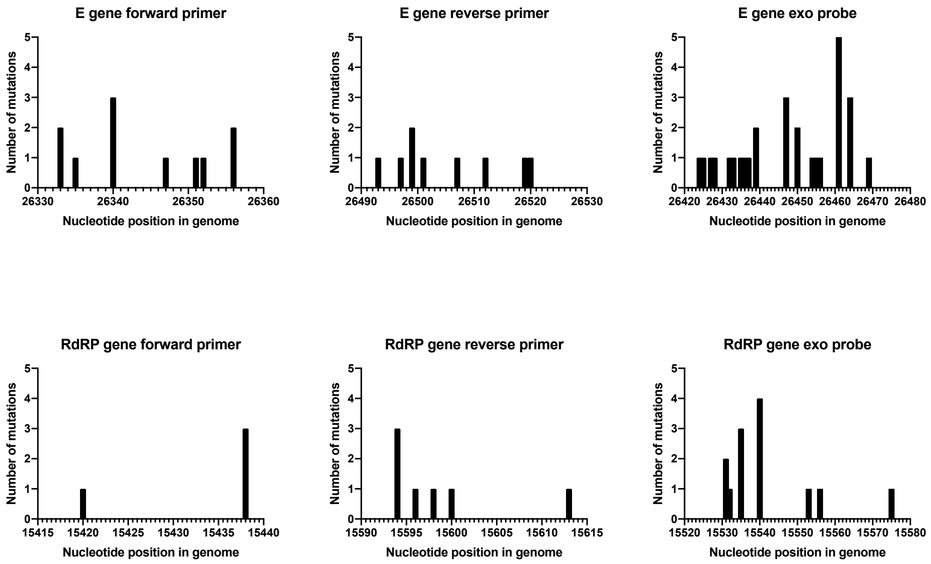


**b**

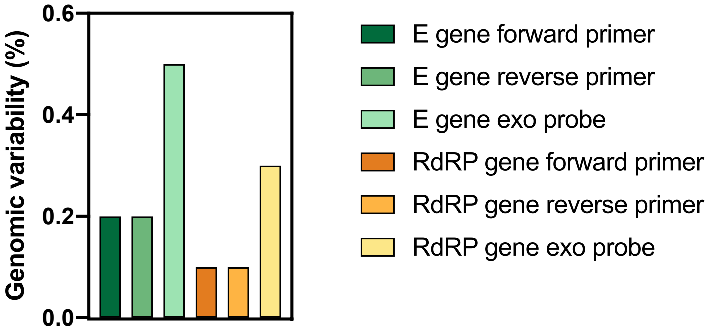


**Supplementary Figure 3.** Analysis of SARS-CoV-2 genomic variations at primers and probes site. (a) The number of mutations was reported for each nucleotide position for the E gene and RdRP primers/probe sets. (b) The overall variability was calculated for the individual primers and probes taking into account the total number of mutations occurring in their sequence. These data came from GISAID genomic epidemiology reports of hCOVID-19 https://www.gisaid.org/epiflu-applications/hcov-19-genomic-epidemiology/ and the mutations were reported from 5139 sequenced genomes (as of October 18$^{th}$, 2020).

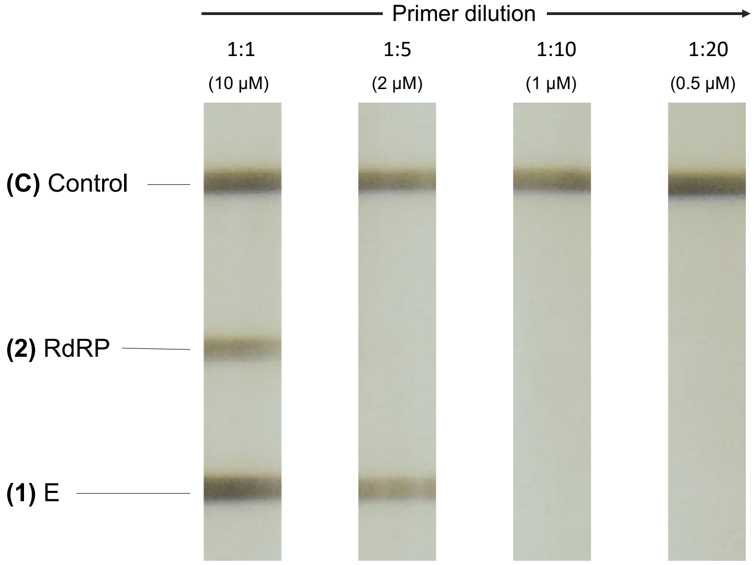


**Supplementary Figure 4.** Photograph of lateral flow tests for optimisation of primer concentration. The modified primers used for detection of the E gene and RdRP gene were diluted to find the optimal concentration which will remove unspecific binding on test lines (1) and (2).

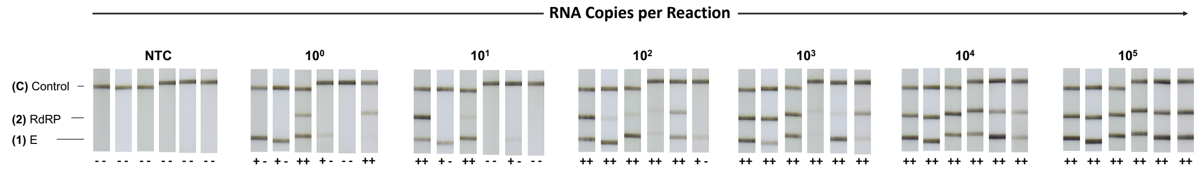


**Supplementary Figure 5.** Photographs of the dipsticks (6 replicates per RNA concentration) used to determine the analytical sensitivity of the dipstick detection method.

**a**

| Target | Primers | Sequence (5’-3’) |
|---|---|---|
| E gene | forward_E_gene (+T7 promoter) | TAATACGACTCACTATAGATGATGAACCGACGACGACT |
| | reverse_E_gene | GATCCGATGCCATGGCTAAAA |
| RdRP gene | forward_RdRP_gene (+T7 promoter) | TAATACGACTCACTATAGTTATGGCCTCACTTGTTCTTGC |
| | reverse_RdRP_gene | AGAGACACTCATAAAGTCTGTGTTG |

**b** **c**

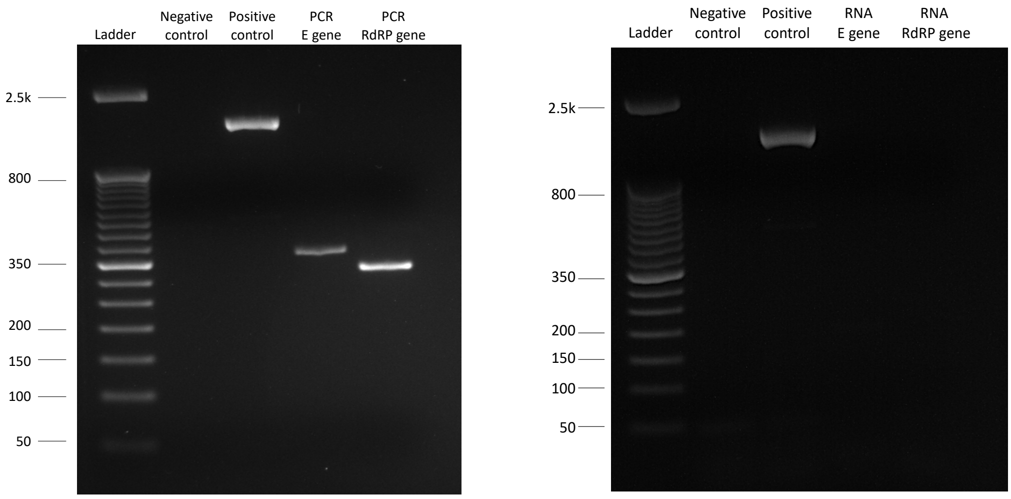


**Supplementary Figure 6.** Primer sequences and agarose gel analysis for the synthesis of RNA standards. (a) Table of the PCR primers used for amplification and addition of T7 promoter for subsequent transcription of SARS-CoV-2 RNA transcripts. (b) Photograph of the agarose gel analysis after amplification of the E gene and RdRP gene by PCR. (c) Photograph of the agarose gel analysis of the transcribed E and RdRP gene RNA transcripts, to verify the purity of the RNA (no DNA impurity).
